# Incidence of and factors associated with SARS-CoV-2 infection among people living with HIV in Southern Spain

**DOI:** 10.1101/2021.03.20.21253397

**Authors:** Marta Fernandez-Fuertes, Anaïs Corma-Gomez, Eva Torres, Elena Rodriguez-Pineda, Ana Fuentes-Lopez, Pilar Rincon, Nieves Fernanddez, Federico Garcia, Samuel Bernal, Luis M Real, Juan Macias, Juan A Pineda

**Affiliations:** Infectious Diseases and Microbiology Unit, Hospital Universitario Virgen de Valme (Sevilla, Spain); Microbiology Service, Hospital Universitario Virgen de Valme (Sevilla, Spain); Microbiology Service, Hospital Universitario San Cecilio (Granada, Spain); Department of Medicine, School of Medicine, University of Sevilla (Sevilla, Spain); Department of Surgery, Biochemistry and Immunology, School of Medicine, University of Malaga (Malaga, Spain)

**Keywords:** HIV, incidence, SARS-CoV-2, COVID-19

## Abstract

**Background:** Whether people living with HIV (PLWH) are at greater risk of acquiring SARS-CoV-2 infection is currently unknown. Prospective serologic studies may allow seroincidence analyses, where all infections are accurately identified. Because of this, we evaluated the incidence of and associated factors with SARS-CoV-2 infection in PLWH in Southern Spain.

**Methods:** This was a prospective cohort study including PLWH from a University Hospital in Southern Spain. Patients were enrolled if 1) they had attended as outpatients our Unit from August 1^st^, 2019 to February 8^th^, 2020; 2) had two subsequent evaluations from February 9^th^, 2020 to February 15^th^, 2021. Serum antibodies against SARS-CoV-2 were determined in baseline and intra-pandemic samples.

**Results:** 710 PLWH were included in the study. Of them, 46 [6.5%, 95% confidence interval (95% CI): 4.8%-8.5%] patients developed SARS-CoV-2 infection. Between May 18^th^ and November 29^th^, 2020, the rate of seroconversion was 5.3% (95% CI: 3.1%-9%) for the general population in the area of Seville and 2.3% (95% CI: 1.3%-3.6%) for PLWH in this study (p=0.001). After multivariate analysis, adjusted by age and sex, active tobacco smoking was the only factor independently associated with lower risk of SARS-Cov-2 infection (Incidence rate ratio 0.35, 95% CI: 0.18-0.68, p=0.002).

**Conclusions:** The incidence of SARS-CoV-2 infection among PLWH in Southern Spain during the ongoing pandemic was lower than that reported for the general population in the same area. Tobacco smoking was the only factor independently associated with a lower risk of incident SARS-CoV-2 infection.

**Summary:** The incidence of SARS-CoV-2 infection among people living with HIV is lower than that of general population in Southern Spain. Active tobacco smoking could be associated with a lower risk of developing COVID-19.

## Introduction

SARS-CoV-2 infection pandemic has extensively involved Spain, with over 2.7 million confirmed cases by January 2021 after three waves. The first reported case in our country was diagnosed in February 9th, 2020 [1]. COVID-19 cases have been observed among people living with HIV (PLWH) in many countries across the world. Several studies suggested that the risk of SARS-CoV-2 infection for PLWH is similar to that of general population [2-4]. However, the evaluation of the incidence of SARS-CoV-2 infection in PLWH have been based in clinical reports. As a substantial part of SARS-CoV-2 infections are asymptomatic [5] and diagnosis was limited by test shortage and health care system capacity surpassing, the incidence of SARS-CoV-2 infection among PLWH could have been underestimated. Thus, whether PLWH are at different risk of acquiring SARS-CoV-2 infection is currently unknown [6].

Similarly, the impact of HIV coinfection on the outcome of SARS-CoV-2 infection has not been clearly defined. PLWH, due to premature ageing and comorbidities [7], could be at a higher risk of poorer COVID-19 outcomes. In fact, after standardization to the age and sex distribution of Spain, the risk of death from COVID-19 was slightly higher among PLWH than for the Spanish general population during the same period [4]. In this same regard, HIV infection was associated with approximately a two-fold increase in COVID-19 mortality in a study from South Africa [2]. In these two studies, the use of tenofovir diphosphate (TDF) plus emtricitabine (FTC) or lamivudine was associated with a lower risk of severe COVID-19 [2, 4]. On the contrary, propensity-matched analyses revealed no difference in COVID-19 outcomes for PLWH, showing that higher mortality is driven by higher burden of comorbidities than in the general population [8]. More importantly, within the same cohort of veterans, HIV infection was not a factor related with outcomes [9].

COVID-19 incidence and outcome data among PLWH based in reported clinical cases, is a limitation for all the aforementioned studies, as milder cases, as well as those in which PCR for SARS-CoV-2 was not carried out, could have gone unnoticed. Serologic studies, in which the diagnosis SARS-CoV-2 infection is based on the detection of serum antibodies, allow prospective seroincidence analyses, where all infections are accurately identified. In this manner, the above-mentioned limitations may be overcome. Because of this, we undertook this study, with the purpose of providing insight on the actual incidence, associated factors and clinical outcome of SARS-CoV-2 infection in PLWH in Southern Spain.

## Patients and Methods

### Design and study patients

This is a study conducted in a cohort of prospectively followed PLWH at the Unit of Infectious Diseases of a university hospital in Seville, Southern Spain. All patients from this cohort are seen at least every six months. At each visit, all patients undergo clinical evaluation and blood drawing for routine testing, as well as sample storage for future determinations. After February 2020, all patients were specifically questioned on respiratory or general symptoms suggesting COVID-19 since the former visit. Serum samples from all patients were cryopreserved at −80°C at each visit. Patients enrolled in that cohort were included in the present study if they fulfilled the following criteria: 1) they had attended the outpatient clinic of our Unit from August 1^st^, 2019 to February 8^th^, 2020 as the day before the first case officially diagnosed in Spain; 2) had two subsequent evaluations from February 9^th^, 2020 to February 15^th^, 2021.

According to data available from the Spanish National Center of Epidemiology (www.cnecovid.isciii.es), the period of study was divided into three periods of time: Baseline or prepandemic period, from August 1^st^, 2019 to February 8^th^, 2020; first pandemic period, corresponding to the first wave in Spain, from February 9^th^, 2020 to May 31^st^, 2020; second pandemic period, corresponding to the second and third pandemic waves in Spain, from June 1^st^, 2020 to February 15^th^, 2021.

### Source of data from general population

For comparisons, data coming from the ENE-COVID survey, a nationwide population-based study to investigate seropositivity for SARS-CoV-2 in the non-institutionalized Spanish population, was used [5]. The study design in this survey includes successive follow-up waves of data collection from random samples of the Spanish population. A first phase with three waves of data collection, from April 27^th^,2020 to June 22^nd^, 2020, and a second phase with one wave of data collection, from November 16^th^ to November 29^th^, 2020, have been completed (https://portalcne.isciii.es/enecovid19/). Data on the incidence of SARS-CoV-2 in the general population living in the province of Seville was gathered from the ENE-COVID study reports (https://portalcne.isciii.es/enecovid19/informes/informe_cuarta_ronda_01.pdf).

### SARS-CoV-2 infection diagnosis

A diagnosis of SARS-CoV-2 infection was established if, at least, one of the following criteria were met: 1) SARS-CoV-2 RNA or antigen was detected in nasopharynx exudate by PCR or immunochromatography, respectively; 2) Seroconversion for SARS-CoV-2 antibodies was documented between the baseline (pre-pandemic) and the follow-up (intra-pandemic) samples.

### Classification of COVID-19 severity

Severe COVID-19 was defined as cases showing at least one of the following data: i) dyspnea, ii) a respiratory rate of 30 or more breaths per minute, iii) a blood oxygen saturation of 93% or less, iv) a ratio of the partial pressure of arterial oxygen to the fraction of inspired oxygen (Pao2:Fio2) of less than 300 mm Hg, or, v) infiltrates in more than 50% of the lung field [10].

### Laboratory procedures

All serum samples were tested for serum SARS-CoV-2 antibodies by electro-chemiluminescence immunoassay (ECLIA), which detects total serum antibodies (including IgG) against this agent (ELECSYS™ Anti-SARS-CoV-2, Roche Diagnostic International, Rotkreutz, Switzerland). The reported sensitivity of this test 14 days after infection is 99.5 % and the specificity for routine diagnostic is 99.8%. PCR analyses for SARS-CoV-2 RNA detection in naso-pharynx exudate were performed by a commercially available procedure (Cobas SARS-CoV-2™ Roche Diagnostic International, Rotkreutz, Switzerland), following the manufacturer instructions.

### Statistical analysis

Continuous variables are expressed as median (Q1–Q3) and categorical variables as numbers (percentage). Cumulative incidences are provided along with 95% confidence interval (95% CI). Frequencies were compared by the chi-square test or the Fisher’s test, when there was at least one cell with an expected frequency lower than 5. The Mann-Whitney U test was used for comparing continuous variables. Differences were considered significant for p values ≤0.05. Poisson regression models were elaborated to assess the factors independently associated with acquisition of SARS-CoV-2 infection. In those analyses, variables related to this condition in the bivariate analysis with a p value <0.2, as well as age and sex, were included. These analyses were carried out using IBM SPSS 25.0 (IBM Corporation) and STATA 12.0 (StataCorp).

### Ethical issues

The study was designed and conducted following the Helsinki declaration. The Ethics committee of the Hospital Universitario Virgen de Valme approved the study. All patients gave written informed consent to be entered in the cohort.

## Results

### Features of the study population

Overall, 710 patients were included in this study. The main baseline features of the study population, as well as the antiretroviral drug combinations they were receiving, are shown in Table 1. Seven hundred and eight individuals (99.7%) were on antiretroviral therapy. Two of 340 patients receiving tenofovir-including regimens were treated with TDF plus FTC. The remaining 340 the patients on tenofovir-based therapy were receiving tenofovir alafenamide (TAF) plus FTC.

**Table 1.** Characteristics of the study population (n=710).

| Parameter | Value |
| --- | --- |
| Age, years<sup>1</sup> | 52 (43-57) |
| Male sex, n (%) | 573 (80.7) |
| HIV infection way, n (%) |  |
| Drug use | 277 (39) |
| Sexual | 387 (54.5) |
| Other and unknown | 46 (6.5) |
| Active tobacco smokers, n (%) | 370 (52.5) |
| Daily alcohol intake $\geq 50$ g, n (%) | 52 (7.4) |
| Active opiate use, n (%) | 48 (6.8) |
| CDC clinical category C, n (%) | 188 (26.5) |
| Nadir CD4 cell counts (cel/uL)<sup>1</sup> | 239 (77-400) |
| Baseline CD4 cell counts (cel/uL)<sup>1</sup> | 633 (412-844) |
| Baseline plasma HIV-RNA $< 50$ c/mL, n (%) | 614 (86.5) |
| ART combination |  |
| TAF or TDF/FTC-based | 340 (48.0) |
| ABC/3TC-based | 97 (13.7) |
| 3TC-including dual therapy | 168 (23.7) |
| Nucleos(t)ide-free | 103 (14.5) |
| Positive plasma HBsAg | 18 (2.6) |
| Positive plasma anti-HCV | 315 (44.5) |
| Active HCV infection | 3 (0.4) |
| Charlson index, n (%) |  |
| 0 | 206 (29) |
| 1-2 | 301 (42.4) |
| 3-5 | 145 (20.4) |
| $\geq 5$ | 58 (8.2) |

1: Median (Q1-Q3).
Abbreviations: ART: Antiretroviral Therapy; TAF: Tenofovir Alafenamide fumarate; TDF: tenofovir disoproxil fumarate; FTC: Emtricitabine; ABC: Abacavir; 3TC: Lamivudine; HBV:

### Incidence of SARS-CoV-2 infection

During the study period, 46 (6.5%, 95% CI: 4.8%-8.5%) patients developed SARS-CoV-2 infection. The diagnosis of SARS-CoV-2 infection was determined by PCR in 17 (37%), by serology in 18 (35%) and by antigen test in 11 (21.7%) of the cases. The frequency of SARS-CoV-2 infection by study period is summarized in Figure 1.

**Figure 1.**
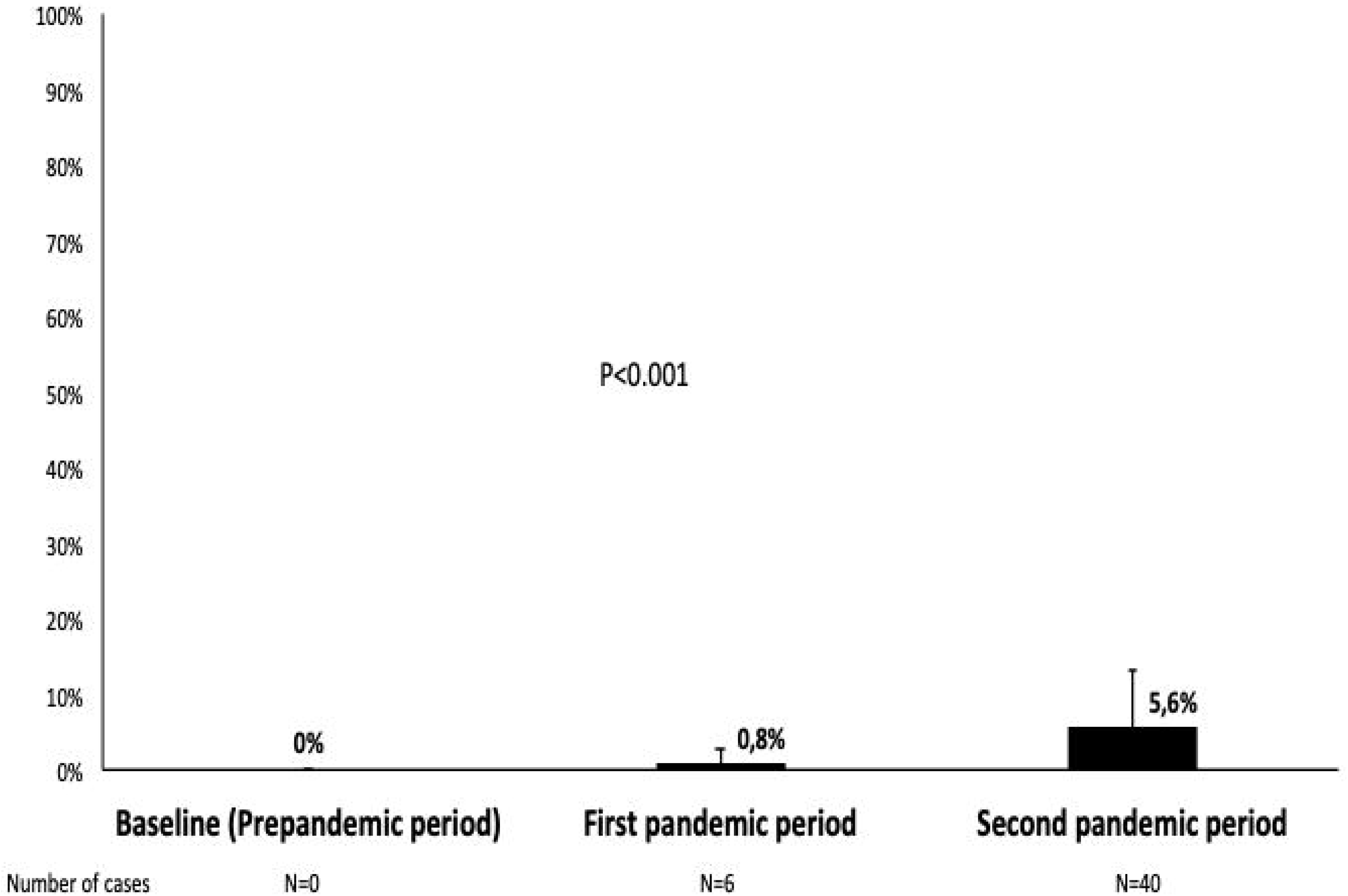
Frequency of SARS-CoV-2 infection by study period.

Overall, 30 (4.3%) of 691 PLWH showed seroconversion during the follow-up. Nineteen (2.3%) patients did not have two follow-up serum samples, six of them because of death during the study period. There were no seroreversions during the study. From May 18^th^, 2020 to November 29^th^, 2020, the rate of seroconversion for the general population in the province of Seville, Southern Spain, was 70 [5.3% (95% CI: 3.1%-9%] of 1326 participants whereas that found among PLWH in this study during the same period of time was 16 [2.3% (95% CI: 1.3%-3.7%)] of 691 patients (p=0.001).

### Clinical outcomes of SARS-CoV-2 infection

Clinical outcomes of SARS-CoV-2-infected patients are shown in Table 2. Five (10.9%) patients were admitted due to severe COVID-19, one (2.2%) of them required Intensive Care Unit admission and subsequently died. Three (60%) patients with severe COVID-19 compared with 3 (7.3%) with asymptomatic or mild disease had previous diagnosis of AIDS (p=0.012). The median (Q1-Q3) nadir CD4 cell counts were 151 (73-279) for patients with severe COVID-19 compared with 332 (88-447) for those without severe COVID-19 (p=0.133). The Charlson index was ≤2 in one (20%) patient with severe COVID-19 vs. 32 (78%) patients with non-severe COVID-19 (p=0.018).

**Table 2.** Clinical outcome of patients with SARS-CoV-2 infection (n=46).

| Clinical presentation | N (%) | Active smokers<br>N (%) | CDC C category<br>N (%) | Nadir CD4<br>counts<sup>1</sup> | Charlson index $\leq 2$<br>N (%) |
| --- | --- | --- | --- | --- | --- |
| Asymptomatic infection | 19 (41.3) | 5 (26) | 2 (11) | 337 (163-448) | 17 (90) |
| Upper-respiratory tract infections | 21 (45.7) | 6 (29) | 1 (4.8) | 311 (62-433) | 14 (67) |
| Extra-respiratory symptoms | 1 (2.2) | 1 (100) | 0 | 514 | 1 (100) |
| Viral pneumonia<sup>2</sup> | 5 (10.9) | 0 | 3 (60) | 138 (46-271) | 1 (20) |

1. Median (Q1-Q3).
2. All five patients with pneumonia required hospital admission, one of them was transferred to the Intensive Care Unit and ultimately died.

Nineteen [41% (95% CI: 3.6%-24%)] PLWH with SARS-CoV-2 infection in this study were asymptomatic, compared with 642 out of 1689 [38% (95% CI: 36%-40%)] among the Seville general population (p=0.650). Regarding patients with severe SARS-CoV-2 infection, needing admission, 16% (95% CI: 14%-18%) patients from the general population were admitted because of severe COVID-19 (n/N=267/1689) compared with 5 [11% (95% CI: 3.6%-24%)] (p=0.363) PLWH in this study.

### Factors associated with incident SARS-CoV-2 infection

Factors associated with SARS-CoV-2 infection are shown in Table 3. HIV way of transmission and smoking tobacco were associated with SARS-CoV-2 infection. Thirty-one (76%) patients with incident SARS-CoV-2 infection were non-smokers at the time of the study. In the multivariate analysis, only non-active tobacco smoking remained independently associated with incident SARS-CoV-2 infection.

**Table 3.**
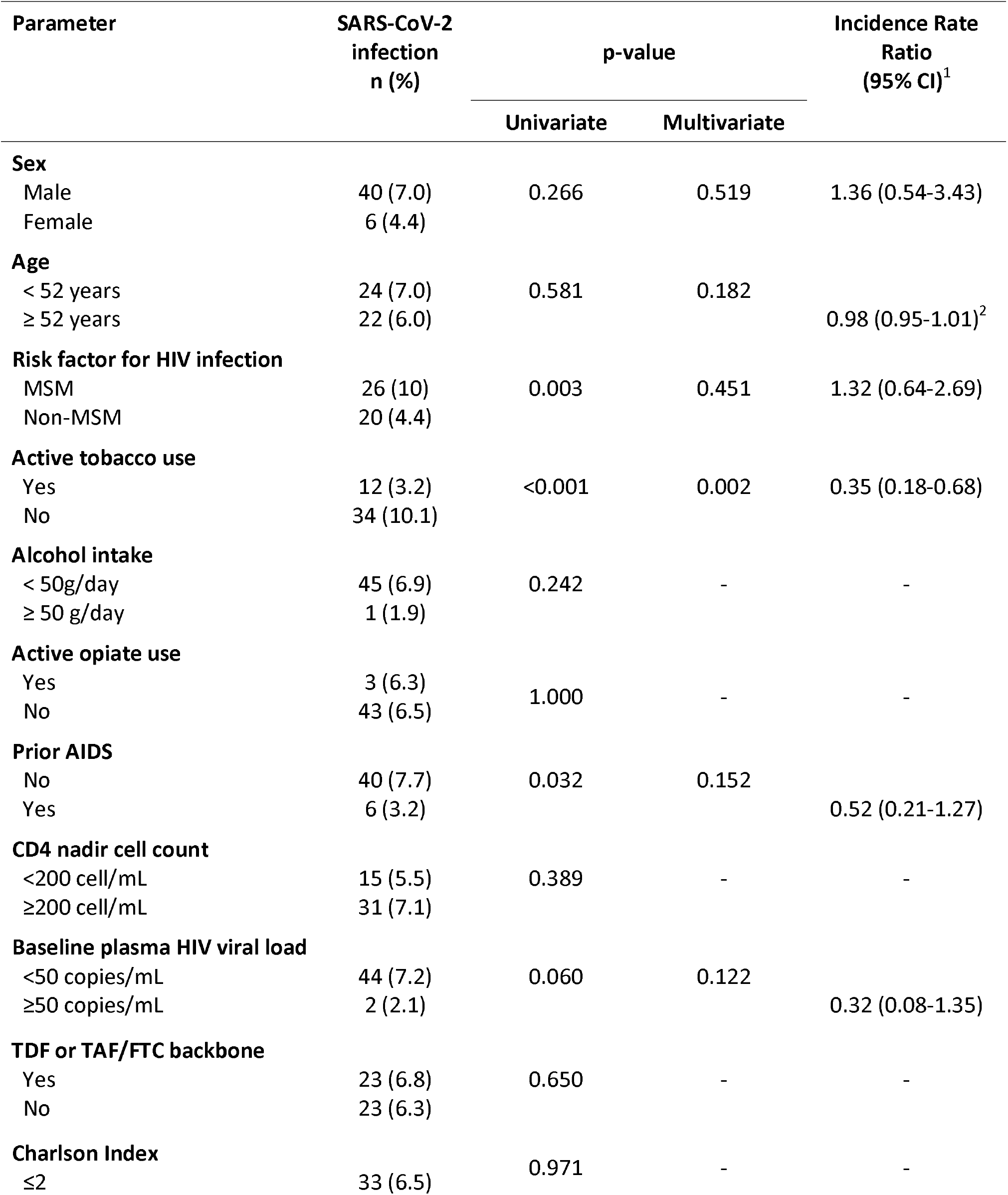
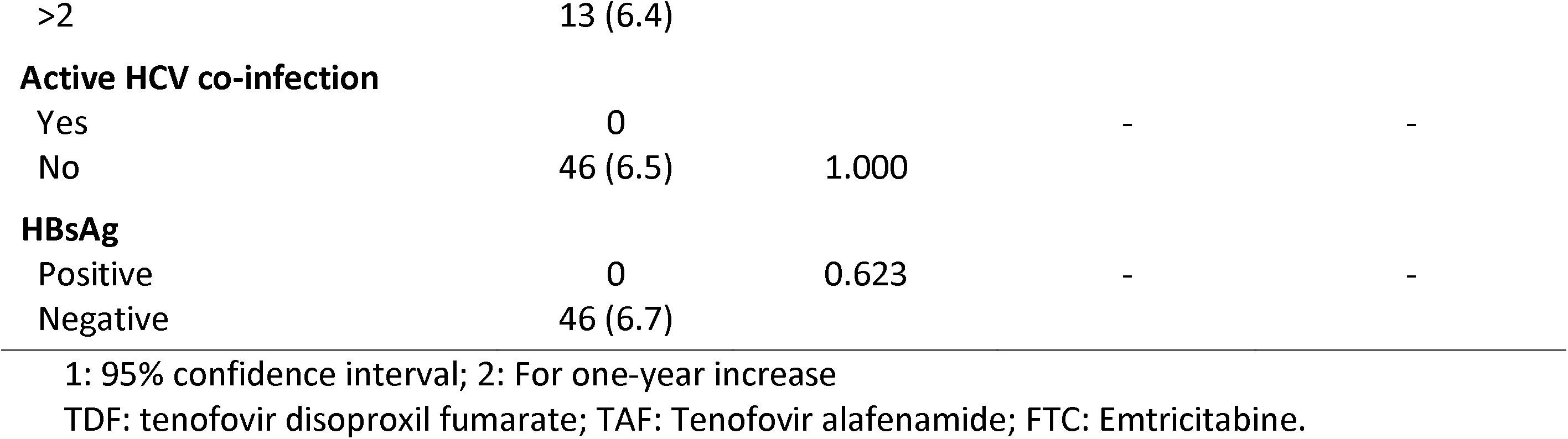
Factors associated with SARS-CoV-2 infection.

The analysis of the relationship between SARS-CoV-2 infection and antiretroviral drugs or drug families did not show any association. Six (6.2%) subjects who were treated with abacavir (ABC) plus lamivudine (3TC)-including combinations, 23 (6.8%) on TDF or TAF/FTC-based regimens, 13 (7.7%) receiving 3TC-including dual therapy and 4 (3.9%) patients on nucleos(t)ide free combinations developed SARS-CoV-2 infection.

## Discussion

The results of this study show that the incidence of SARS-CoV-2 infection among PLWH in Southern Spain during the ongoing pandemic was lower than that reported for the general population in the same area. Tobacco smoking was the only factor independently associated with the risk of incident SARS-CoV-2 infection. Thus, active smokers were at a lower risk of SARS-CoV-2 infection than non-active smokers.

The seroincidence of SARS-CoV-2 infection was significantly lower for PLWH than for the general population living in the province of Seville, Southern Spain. Three quarters of the participants in the first phase, conducted after the first wave in Spain, were also included in the second phase survey in the ENE-COVID study (https://portalcne.isciii.es/enecovid19/informes/informe_cuarta_ronda.pdf). This provided fairly precise data on seroconversions among general population during the follow-up waves, specifically between the first and second phases. We tested a prospective cohort of PLWH for antibodies against SARS-CoV-2 before the onset of the pandemic and during the pandemic. For comparisons, we evaluated seroconversions in the cohort selecting the same initial and final dates for data collection in the ENE-COVID survey. Thus, the differences in SARS-CoV-2 infection seroincidence between PLWH and the general population reported herein can be regarded as accurate.

The reasons why the incidence of SARS-CoV-2 among PLWH is lower than in the general population in our area are unclear. Theoretically, this fact could have, at least, two explanations. First, that antiretroviral therapy decreases the risk of acquiring SARS-CoV-2 infection. In our opinion, this is unlikely, as no association between the risk of SARS-CoV-2 and different antiretroviral combinations were found in the study population. The second reason could be that PLWH in our area are more aware of the risks associated with COVID-19 that a great part of the general population and, consequently, were more compliant with general measures for the prevention of SARS-CoV-2 infection, such as mask wearing, social distance and hand hygiene. Given that, PLWH included here are used to be very compliant with medical recommendations, we believe that the latter is the most conceivable reason underlying the lower incidence of SARS-CoV-2 infection in this population.

In our study, SARS-CoV-2 infection in PLWH was not significantly more severe than in the general population recruited in the ENE-COVID survey. The rates of asymptomatic infection were similar, as were the proportion of individuals with severe COVID-19. We cannot rule out that some patient who had developed asymptomatic infection at the beginning of the pandemic tested negative for SARS-CoV-2 serology latter, because of plasma antibody vanishing. However, plasma antibodies against SARS-CoV-2 disappear in very few asymptomatic patients [11], making this potential underdiagnosis of asymptomatic infections very unlikely. Indeed, antibodies do not decline in the short-term after diagnosis [12]. In our study, we found no case of serorreversion. Contrary to other reports in PLWH [13-15], COVID-19 was not more severe in HIV infection compared with HIV-uninfected persons. In our study COVID-19 severity depended on the Charlson index, compounded of comorbidities and age. Morover, severe COVID-19 in PLWH in our study was also associated with previous AIDS diagnosis. Nadir CD4 cell counts were numerically lower for patients with severe COVID-19, but statistical significance was not reached. Immunosuppression, i.e. current CD4 cell counts, was also associated with COVID-19 outcomes in a previous study [16]. Thus, in addition to age and comorbidities, historical immunosuppression, as reflected both by previous AIDS diagnosis and nadir CD4 cell counts, should be taken into account for COVID-19 risk stratification among PLWH.

Surprisingly, active tobacco smoking was independently associated with a lower incidence of SARS-CoV-2 infection. However, this finding is in agreement with a recent report on a COVID-19 outbreak on a French Navy nuclear aircraft carrier [17]. Among crewmembers, current smoking was associated with a lower risk of developing SARS-CoV-2 infection. Smoking is characterized by inhalation and by repetitive hand-to-mouth movements, which may increase the chances of viral contamination. Moreover, nicotine induces angiotensin-converting enzyme II (ACE-2) overexpression in human bronchial epithelial cells [18]. Because of these, and increased incidence and/or severity of COVID-19 could be expected. The underlying mechanisms involved in this paradoxical and apparent protective effect of tobacco smoking need to be clarified in further studies.

In this study, no association between antiretroviral drugs or combinations and the risk for SARS-CoV-2 infection was found. Previous studies have shown that TDF has potent antiviral effect against SARS-CoV-2, because it tightly binds the viral RNA-dependent RNA polymerase [19]. Because of this, clinical trials aimed to assess the efficacy of TDF, a tenofovir salt which reaches higher plasma concentration than TAF, both in the prevention and treatment of COVID-19 were undertaken [20]. Furthermore, two retrospective studies suggest an association between TDF plus FTC treatment and the prevention of SARS-CoV-2 acquisition, hospitalization or death associated with COVID-19 [2, 4]. In our study, only two patients were on TDF plus FTC, because most individuals in this cohort had been switched from TDF to TAF in the last few years. Consequently, we were unable to analyze the effect of TDF on the risk of SARS-CoV-2 infection.

This work may have some limitations. First, incidence comparisons were not age nor sex-standardized. PLWH from this cohort are relatively younger, could bear less comorbidities than the general population and includes a higher proportion of males. Nevertheless, the analysis of factors associated with incident SARS-Cov-2 infection did not show the risk of infection varied according to sex or Charlson index, which includes age and comorbidities. Second, the study results are applicable to settings where most patients are under effective antiretroviral therapy, with HIV replication suppression and high CD4 cell counts. The present study conclusions may not be valid for PLWH in advanced or untreated HIV infection. As a counterpart, this was a prospective cohort study, with pre-planned visits and scheduled sera collection, which allows precisely estimating incident SARS-CoV-2 infection and the outcome of COVID-19. Since prospective data on the incidence and risk of SARS-CoV-2 infection in PLWH are lacking [6], the present study fills part of this gap. These are the most important strengths of this study.

In summary, the incidence of COVID-19 among PLWH in Seville, Southern Spain after three waves of COVID-19 pandemic was lower to that observed in the general population in the same geographical area. The severity of COVID-19 was similar to that in patients without HIV infection and determined by comorbidities and age. Nevertheless, some HIV-related factors, as historical immunosuppression, could influence the outcome of COVID-19. The apparent protector role of tobacco smoking on the risk of SARS-CoV-2 infection deserves further evaluation, as it might lead to open ways to develop drugs for COVID-19 treatment or prophylaxis.

## Data Availability

There is no other data

